# Cross-site predictions of readmission after psychiatric hospitalization with mood or psychotic disorders

**DOI:** 10.1101/2024.08.26.24312586

**Authors:** Boyu Ren, WonJin Yoon, Spencer Thomas, Guergana Savova, Timothy Miller, Mei-Hua Hall

**Affiliations:** Laboratory for Psychiatric Biostatistics, McLean Hospital, Belmont, MA; Computational Health Informatics Program, Boston Children’s Hospital, Boston, MA; Department of Pediatrics, Harvard Medical School, Boston, MA; Psychosis Neurobiology Laboratory, McLean Hospital, Belmont, MA; Department of Psychiatry, Harvard Medical School, Boston, MA

## Abstract

Patients with mood or psychotic disorders have high rates of unplanned readmission, and predicting readmission likelihood may guide discharge decisions. In this retrospective, multi-site study, we assess the predictive power of various structured variables from electronic health records for all-cause readmission in each site separately and evaluate the generalizability of the in-site prediction models across sites. We find that the set of relevant predictors vary significantly across. For example, length of stay is strongly predictive of readmission at only three out of the four sites. We also find a general lack of cross-site generalizability of the in-site prediction models, with in-site predictions having an average F1 score of 0.666, compared to an average F1 score of 0.551 for cross-site predictions. The generalizability cannot be improved even after adjusting for differences in the distributions of predictors. These results indicate that, with this set of predictors, fitting individual models at each site is necessary to achieve reasonable prediction accuracy. Additionally, they suggest that more sophisticated predictors variables or predictive algorithms are needed to develop generalizable models capable of extracting robust insights into the root causes of early psychiatric readmissions.

## Introduction

Mood and psychotic disorders rank among the most disabling conditions worldwide.^1–5^ In 2020, the United States alone spent $186 billion on treatment for mental health disorders, constituting 6.4% of the total US healthcare spending.^6^ This underscores the substantial economic burden of mental health conditions on the healthcare system. In addition, a substantial proportion of psychiatric inpatients are readmitted unplanned within 30 days after discharge.^7,8^ Readmissions not only are disruptive but also cause enormous economic burden for patients and families, and are a key driver of rising healthcare costs.^9,10^ The readmission rate is considered an indicator of the mental health care quality.^11,12^ A 2013 US Nationwide Readmission study reported that psychiatric disorders account for the largest portion (24%) of short-term (< 30-day) readmissions among young adults between 18 and 44 years old; that the readmission rate of schizophrenia patients is among the highest (25.7%); and that psychiatric disorders, including substance abuse, contribute to a tenth of all discharges leading to readmission, with nearly one fifth of discharges leading to readmission occurring in Medicare and Medicaid insured individuals.^13^

Addressing and predicting unplanned readmission are therefore major unmet needs of psychiatric care.

Previous studies leveraging structured elements of Electronic Health Record (EHR) data for short-term psychiatric readmission prediction have identified various risk factors associated with 30-day unplanned psychiatric readmissions, including gender, marital status, insurance status, specific diagnoses (e.g., schizophrenia), substance use, and length of stay.^7,14^ More recently, Natural Language Processing (NLP) techniques have been employed to extract insights from unstructured clinical notes for readmission risk prediction. These studies corroborated findings from structured elements while uncovering additional features such as suicidality,^15^ prior psychiatric admissions,^16^ family relationships,^16,17^ and symptom severity (e.g., mood, psychosis).^16–18^

Despite the potential of EHR data in readmission prediction, several limitations persist. One important issue is that there is a scarcity of cross-dataset validation. Most studies focus on model development and validation within a single hospital or dataset drawn from a large healthcare system consisting of different hospitals. For the latter, it is assumed that hospital-specific differences in practices, documentation, and patient populations within the same healthcare system are largely negligible. However, each hospital might have a history of catering to specific patient populations and psychiatric needs. True cross-dataset validation, where a model trained on one hospital dataset is explicitly tested on another, is exceedingly rare. This limitation hampers our understanding of how well prediction models and predictive variables generalize across different hospitals and patient populations. Furthermore, direct comparisons of various prediction algorithms across independent datasets are uncommon, limiting our understanding of how models might perform in diverse settings with various patient populations or institutional patient care practices. These issues are important considerations for building robust and widely applicable readmission prediction models.

To address these issues, we extract psychiatric inpatient EHR from four different hospitals within the same healthcare system in the Boston area. We focus on assessing characteristics of patient populations across hospitals, to perform cross-dataset validation, where a model trained on one hospital dataset is tested on another, and to compare various prediction algorithms across independent hospital datasets.

## Materials and methods

### Study design and cohort generation

This is a retrospective study of patients with electronic health records (EHR) of inpatient psychiatric unit stays at four Boston academic medical hospitals within the Mass General Brigham (MGB) system – Massachusetts General Hospital (MGH), McLean Hospital (MCL), Brigham and Women’s Hospital (BWH), and Faulkner Hospital (FH). EHRs were sourced from the Research Patient Data Registry (RPDR), a centralized regional data repository across all institutions in the MGB healthcare system. While these four hospitals have different patient populations, staff, and operate largely independently, their records are all accessible in the RPDR research database system. This study was approved by the MGB Institutional Review Board.

We queried EHRs of patients, separate in each hospital, who were age between 18-65 years old at the time of query (April 2023) AND with any diagnosis codes for Mood disorders (F30-F39 in ICD10) OR psychotic disorders (F20-29 in ICD10). While ICD codes may have low positive predictive value for phenotyping studies,^19^ for this work we took a liberal view of the inclusion criteria in order to take a broader view of these disorders, as well as to increase the size of our labeled datasets. Structured data included demographics (age, sex, race/ethnicity, insurance type, and diagnoses). Data also included unstructured clinical narratives (progress reports and discharge summaries), analysis of which will be described in future work.

### Data Extraction

To study all-cause readmission, we extracted the set of patients with inpatient encounters matching the above inclusion criteria, and then all additional inpatient encounters for those patients, across MGB sites. The i2b2^20^ query interface at MGB allows us to specify these criteria in a graphical user interface. The query results were returned as text files representing database tables, with one row per encounter. To create classification instances, we automatically labeled inpatient encounters as being “psychosis or mood disorder related” if they contained an ICD10 billing code starting with F2 or F3. We then collected the set of additional encounters for these patients and ordered all encounters by date. The database column contains an “Inpatient/Outpatient” column which we use to define Inpatient encounters. Inpatient encounters that overlapped in dates were merged during pre-processing. These merged inpatient encounters represent the classification instances. For each classification instance, it was given a positive label (representing early readmission) if the admission date of the next inpatient encounter in the patient encounter sequence was within 30 days of the discharge date of the discharge encounter.

### Prediction algorithms and variables

We considered two models for the prediction of 30-day readmission, logistic regression and random forest (RF). Logistic regression is the most widely used classification model with a linear decision boundary while RF allows for a non-linear decision boundary and the ability to handle complex interactions between predictors. For both models, we used age at discharge, sex, race, diagnosis of psychosis, mood disorder, anxiety and substance use disorder (SUD), and length of stay (in days) as the predictors. We only included main effects of the predictors in the logistic regression model for simplicity. We then evaluated the within-site and cross-site prediction accuracy of these models. For each site, we split the original data with a 3-to-1 ratio into a training and a testing set. To create these splits, we use the last digit of a cross-site patient identifier so that patients with encounters at more than one site are in the same split at different sites (i.e., a patient would not be in the train set at McLean and the test set at MGB). We developed a site-specific model using the training set for a given site and applied these models to the testing set of the same site for within-site evaluation and the testing set of other sites for cross-site evaluation. We used the area under the ROC curve (AUROC) and F1 score to measure the prediction performance.

### Cross-site prediction with covariate shift

The site-specific prediction model for site *s* is designed to optimize the expected prediction performance with respect to the data distribution of site *s*. When applying it to a different site, it might not be able to produce useful predictions if the data distribution changes. With the aim to improve the cross-site prediction performance, we used a simple reweighting strategy to account for a particular type of shift in data distributions across sites, covariate shift, which assumes that the only difference in data distributions is in the marginal distributions of the predictors while the conditional distribution of the outcome given the predictors remain identical across sites [21]. This method works by weighting each of the input samples at a source site, when training a prediction model aiming to transport information from the source study to a target study, such that the weighted distribution of the predictors matches the distribution at the target site. For example, if the target site has a larger proportion of females than the source site, the weighting scheme would up-weight female subjects and down-weight males in model training. For a more formal description of this method, see Appendix A.

## Results

### Summary statistics of the dataset

Table 1 shows a detailed breakdown of psychiatric inpatient encounters (i.e., classification instances), their breakdown across sites, and the statistics of the variables associated with them. The initial query and post-processing yielded *n*=52237 classification instances across four sites: MCL (n=29845), MGH (n=18772), BWH (n=1406), FH (n=1053). From the table we can find that no pair of sites share similar distributions across all predictors. Clustering structure is evident for certain predictors. For example, records in BWH and Faulkner have similar racial composition while MCL and MGH share the same racial distribution. However, the clustering structure varies across different predictors. A patient may have multiple F2 and/or F3 diagnoses in a single encounter, most of the encounters have a diagnosis of mood disorders (lowest prevalence is 75.2% in MCL). Diagnosis of psychosis, anxiety and SUD are less common. The proportion of 30-day readmission is not too extreme in all four sites, ranging from 21.1% in Faulkner to 43.3% in MGH.

**Table 1:**
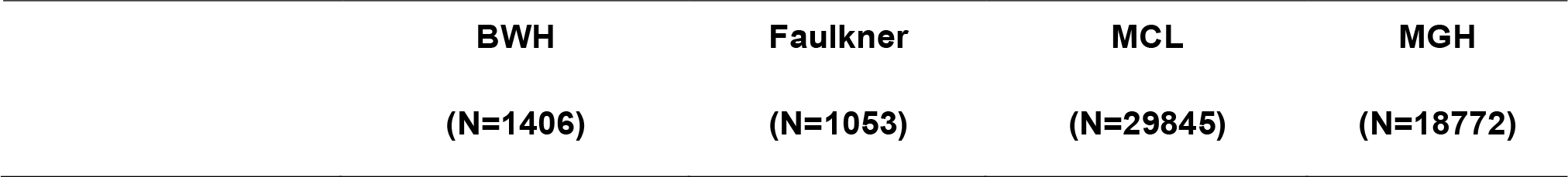
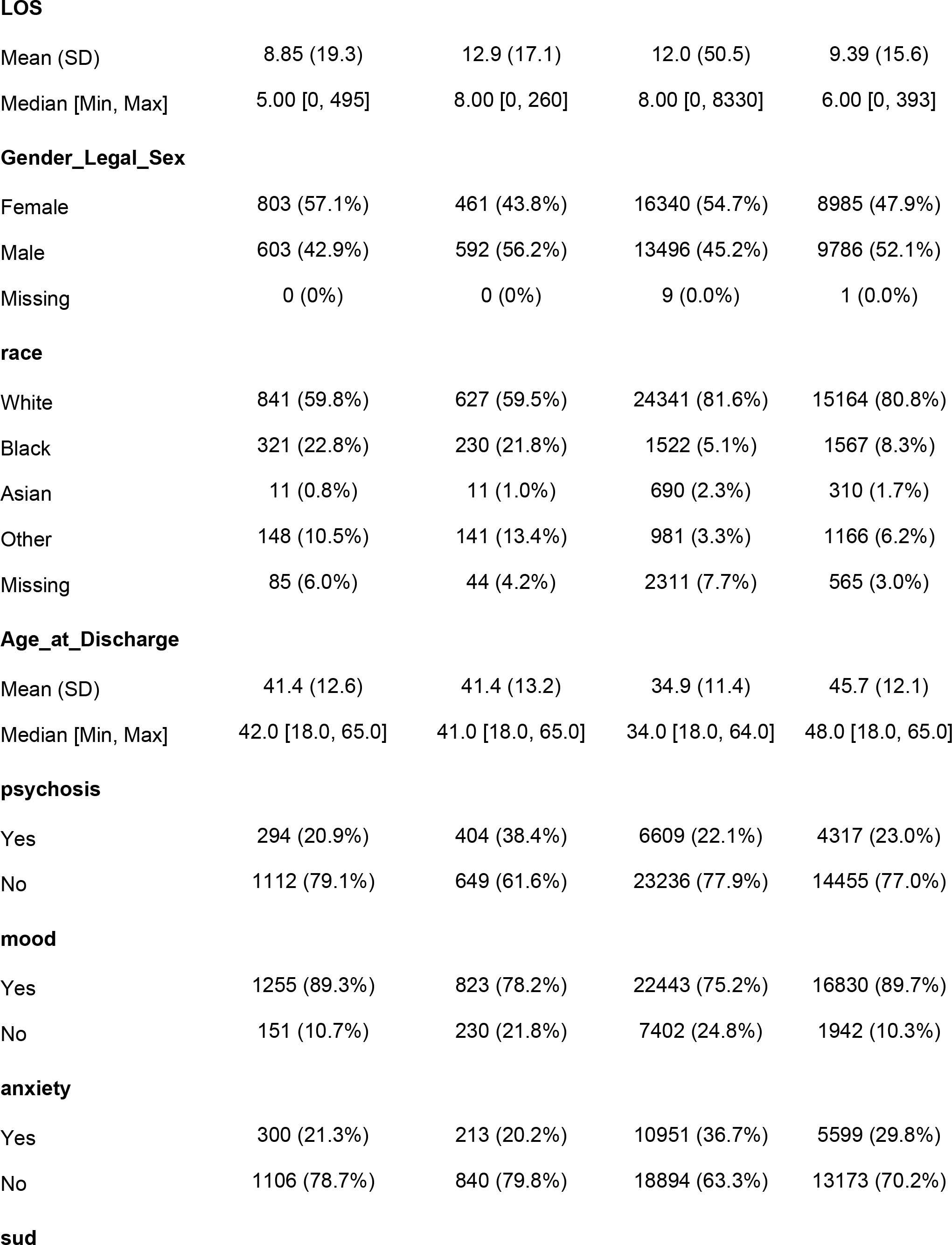
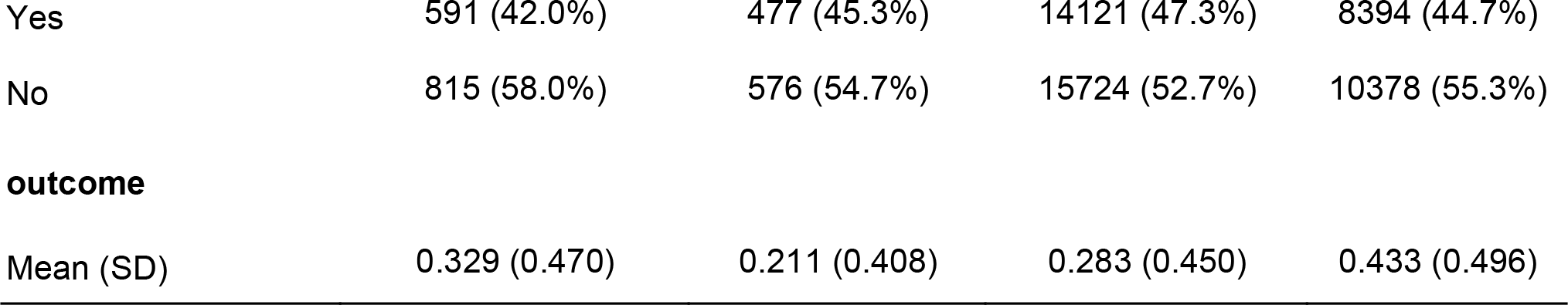
Distributions of predictors in consideration and the outcome (30-day readmission) in four study sites. BWH=Brigham and Women’s Hospital, MCL=McLean, MGH=Mass General Hospital. LOS: length of stay; sud: substance use disorder

| BWH | Faulkner | MCL | MGH |
| --- | --- | --- | --- |
| (N=1406) | (N=1053) | (N=29845) | (N=18772) |

**LOS**
|  |  |  |  |  |
| --- | --- | --- | --- | --- |
| Mean (SD) | 8.85 (19.3) | 12.9 (17.1) | 12.0 (50.5) | 9.39 (15.6) |
| Median [Min, Max] | 5.00 [0, 495] | 8.00 [0, 260] | 8.00 [0, 8330] | 6.00 [0, 393] |

**Gender\_Legal\_Sex**
|  |  |  |  |  |
| --- | --- | --- | --- | --- |
| Female | 803 (57.1%) | 461 (43.8%) | 16340 (54.7%) | 8985 (47.9%) |
| Male | 603 (42.9%) | 592 (56.2%) | 13496 (45.2%) | 9786 (52.1%) |
| Missing | 0 (0%) | 0 (0%) | 9 (0.0%) | 1 (0.0%) |

**race**
|  |  |  |  |  |
| --- | --- | --- | --- | --- |
| White | 841 (59.8%) | 627 (59.5%) | 24341 (81.6%) | 15164 (80.8%) |
| Black | 321 (22.8%) | 230 (21.8%) | 1522 (5.1%) | 1567 (8.3%) |
| Asian | 11 (0.8%) | 11 (1.0%) | 690 (2.3%) | 310 (1.7%) |
| Other | 148 (10.5%) | 141 (13.4%) | 981 (3.3%) | 1166 (6.2%) |
| Missing | 85 (6.0%) | 44 (4.2%) | 2311 (7.7%) | 565 (3.0%) |

**Age\_at\_Discharge**
|  |  |  |  |  |
| --- | --- | --- | --- | --- |
| Mean (SD) | 41.4 (12.6) | 41.4 (13.2) | 34.9 (11.4) | 45.7 (12.1) |
| Median [Min, Max] | 42.0 [18.0, 65.0] | 41.0 [18.0, 65.0] | 34.0 [18.0, 64.0] | 48.0 [18.0, 65.0] |

**psychosis**
|  |  |  |  |  |
| --- | --- | --- | --- | --- |
| Yes | 294 (20.9%) | 404 (38.4%) | 6609 (22.1%) | 4317 (23.0%) |
| No | 1112 (79.1%) | 649 (61.6%) | 23236 (77.9%) | 14455 (77.0%) |

**mood**
|  |  |  |  |  |
| --- | --- | --- | --- | --- |
| Yes | 1255 (89.3%) | 823 (78.2%) | 22443 (75.2%) | 16830 (89.7%) |
| No | 151 (10.7%) | 230 (21.8%) | 7402 (24.8%) | 1942 (10.3%) |

**anxiety**
|  |  |  |  |  |
| --- | --- | --- | --- | --- |
| Yes | 300 (21.3%) | 213 (20.2%) | 10951 (36.7%) | 5599 (29.8%) |
| No | 1106 (78.7%) | 840 (79.8%) | 18894 (63.3%) | 13173 (70.2%) |

**sud**
|  |  |  |  |  |
| --- | --- | --- | --- | --- |
| Yes | 591 (42.0%) | 477 (45.3%) | 14121 (47.3%) | 8394 (44.7%) |
| No | 815 (58.0%) | 576 (54.7%) | 15724 (52.7%) | 10378 (55.3%) |

|  |  |  |  |  |
| --- | --- | --- | --- | --- |
| Mean (SD) | 0.329 (0.470) | 0.211 (0.408) | 0.283 (0.450) | 0.433 (0.496) |

### In-site Predictions

The estimated regression coefficients of the logistic model for all sites are illustrated in Table 2. None of the predictors are consistently significant across all sites and the estimated odds ratio can be qualitatively different from site to site. Length-of-stay (LOS) is significantly associated with the outcome in three sites but with different directionality (negative association in MGH and positive associations in BWH and Faulkner). Sex, diagnosis of anxiety and SUD are significantly associated with the outcome in MGH and MCL, although the directionality of the association of diagnosis of anxiety is different between these two sites. Age at discharge only shows significance in BWH and the contrast between other races to white, diagnosis of psychosis and mood disorders only show significance in MGH.

**Table 2:** Estimated regression coefficients of the logistic models for each of the four sites. Note the regression coefficients are transformed into the scale of odds ratios.

|  | MGH |  | MCL |  | BWH |  | FH |  |
| --- | --- | --- | --- | --- | --- | --- | --- | --- |
| *Predictors* | *Odds Ratios* | *p* | *Odds Ratios* | *p* | *Odds Ratios* | *p* | *Odds Ratios* | *p* |
| Age at Discharge | 1 | 0.245 | 1 | 0.26 | 0.99 | **0.015** | 1 | 0.701 |
| Gender Legal Sex [Male] | 1.4 | **<0.001** | 1.11 | **0.002** | 1.13 | 0.403 | 1.02 | 0.918 |
| race [Black] | 0.95 | 0.445 | 0.94 | 0.36 | 1.2 | 0.285 | 1.06 | 0.81 |
| race [Asian] | 0.81 | 0.152 | 1.09 | 0.395 | 1.5 | 0.552 | 2.17 | 0.285 |
| race [Other] | 0.78 | **0.001** | 0.95 | 0.544 | 1.31 | 0.232 | 0.91 | 0.755 |
| psychosisTRUE | 0.49 | **<0.001** | 0.92 | 0.065 | 1.06 | 0.815 | 1.44 | 0.156 |
| moodTRUE | 0.61 | **<0.001** | 1.02 | 0.578 | 1.05 | 0.885 | 1.11 | 0.717 |
| anxietyTRUE | 0.63 | **<0.001** | 1.09 | **0.01** | 0.77 | 0.154 | 1.28 | 0.28 |
| sudTRUE | 0.39 | **<0.001** | 0.62 | **<0.001** | 1.1 | 0.519 | 1.01 | 0.962 |
| LOS | 0.98 | **<0.001** | 1 | 0.341 | 1.01 | **0.007** | 1.01 | **0.039** |

The results of the feature importance study using RF are shown in Table 3. The feature importance, measured by the decrease in prediction accuracy when randomly permuting a variable while keeping all other variables intact, reveals a different landscape of predictive power associated with each predictor. For all four sites, LOS has the highest importance among all predictors. However, the actual magnitude of this predictor is quite small except in the case of MGH (0.175). All other predictors are much less predictive, with the highest importance achieved for substance use disorder (sud) at 0.016. Note that the accuracy of a prediction model ranges from 0 to 1. A decrease of more than 0.1 can be considered as a substantial drop in the model performance while a decrease less than 0.01 is likely negligible.

**Table 3:**
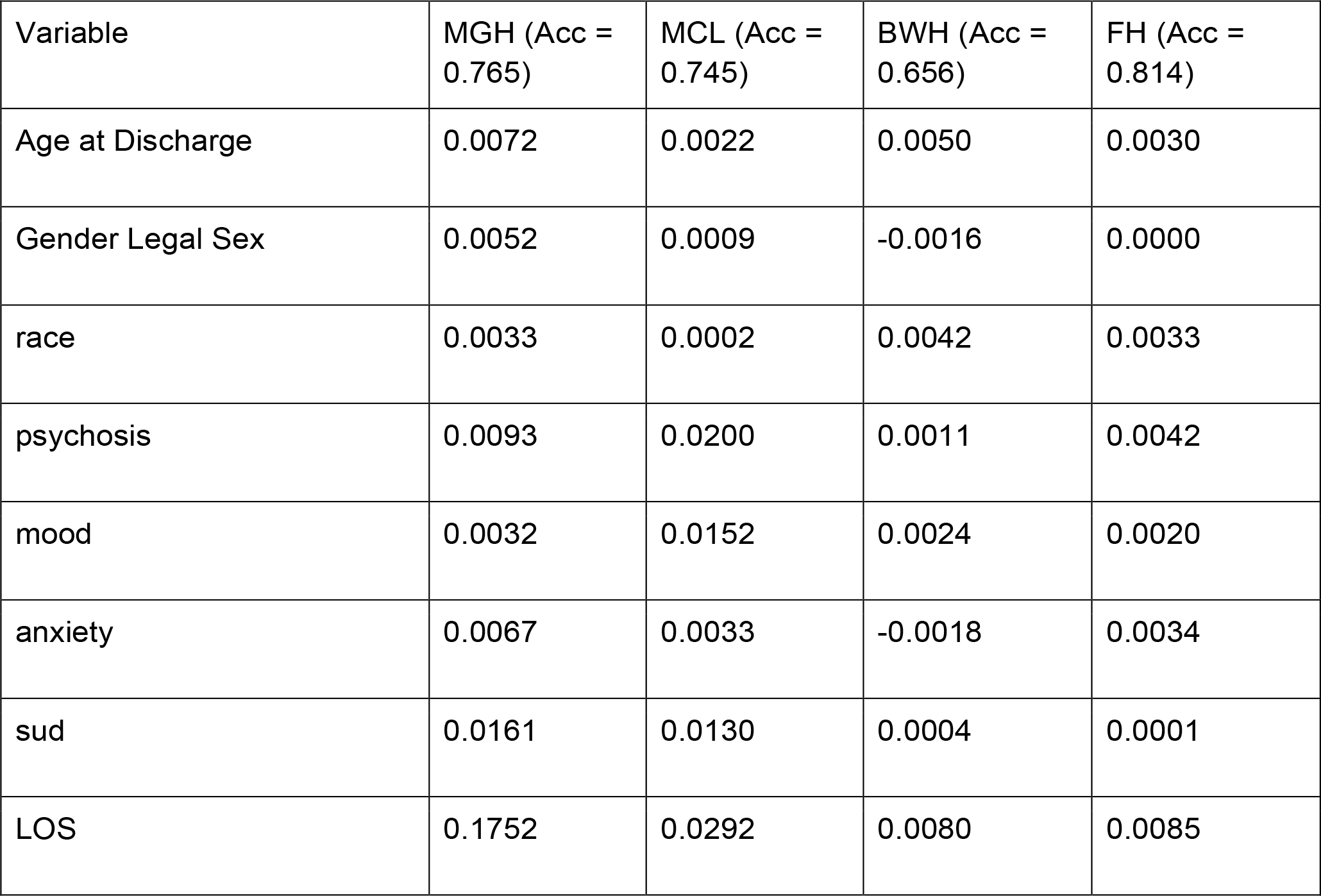
Feature importance of all predictors, measured by amount of accuracy decrease when removing the variable from the RF, over all four sites. Larger positive values means higher importance. “Acc” indicates the out-of-bag estimate of accuracy of the model with all variables included.

The AUCs and F1 score of the logistic models applied to the testing data of the same sites are visualized in the top panels of Figure 1 (diagonal elements of the heatmaps). Except for MGH, none of the sites achieve an AUC larger than 0.6, indicating low level of discriminative power of these models. MGH also has the highest F1 score among all sites. The results for RF are shown in the bottom panels of Figure 1. Only MGH enjoys a discernible improvement in prediction performance when switching from logistic model to RF—the AUC for MGH increases from 0.687 to 0.738 and the F1 score from 0.68 to 0.749. All results combined suggest that the predictors we included are only suitable for the prediction of 30-day readmission for encounters in MGH.

**Figure 1:**
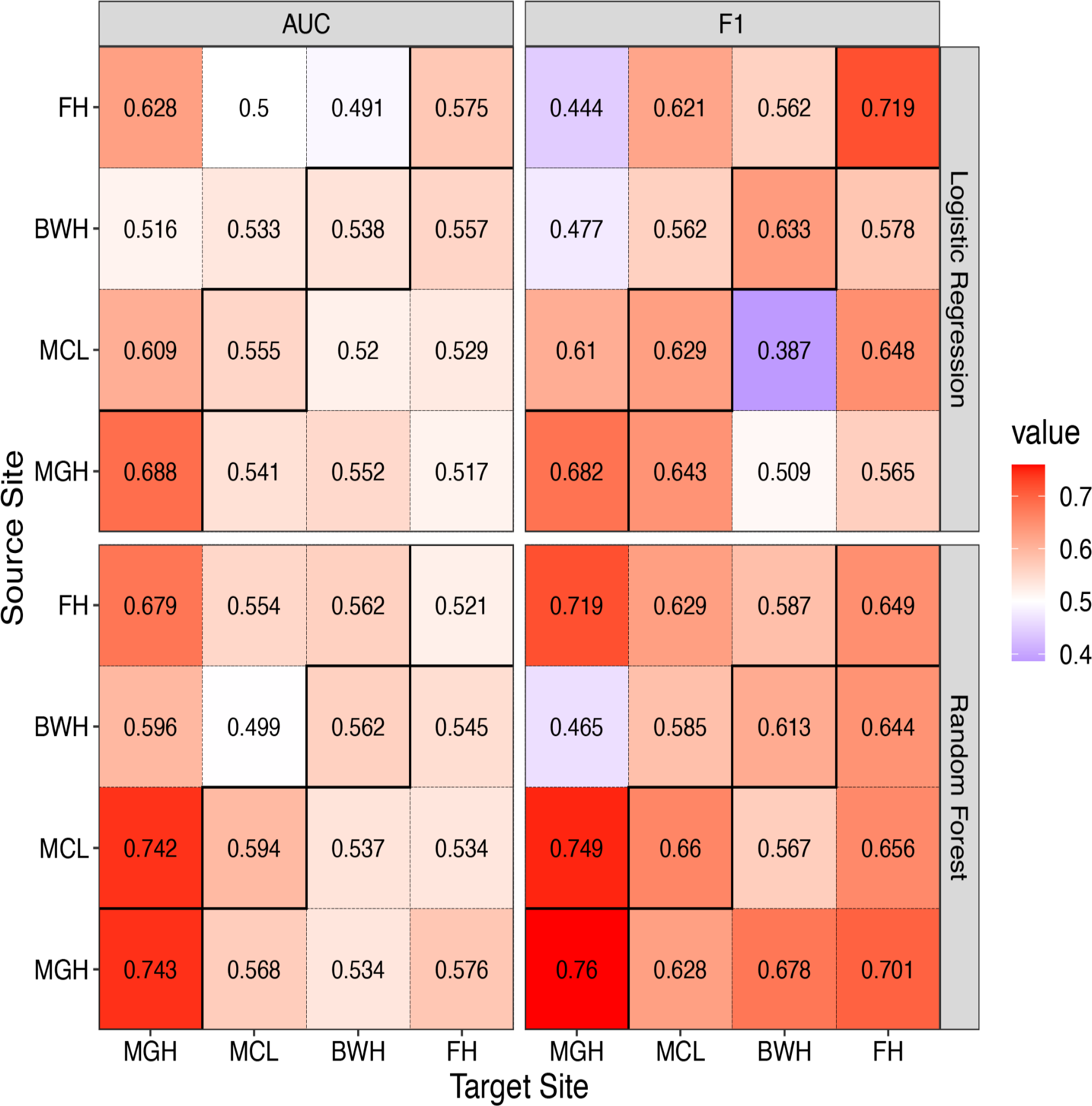
In- and cross-site prediction performance of the logistic regression models (top) and RF (bottom). The tiles with solid borders indicate the in-site prediction performance.

### Cross-site Predictions

When directly applying a prediction model trained on data from one site to testing data from another site, the prediction performance is generally worse than in-site predictions (see Figure 1, off-diagonal elements), demonstrating a general lack of transportability of the prediction models across sites. The only exception is MGH. Faulkner and MCL models both achieve reasonable AUCs and F1-scores when tested on MGH data. RF trained with MCL data even leads to a higher AUC than in-site predictions for MGH. This indicates that the MGH records are inherently easier to predict.

One potential reason for this observed lack of transportability is due to covariate shift, that is, the distribution of the predictors are different from site to site, which has been confirmed in Table 1. The performance of cross-site predictions that adjust for covariate shift, based on the IPW approach introduced in the Method section, is illustrated in Figure 2. The results clearly show that the adjustment for covariate shift has relatively low benefit to transportability of the prediction models, as nearly none of the AUCs or F1 scores were improved substantially after weighting.

**Figure 2:**
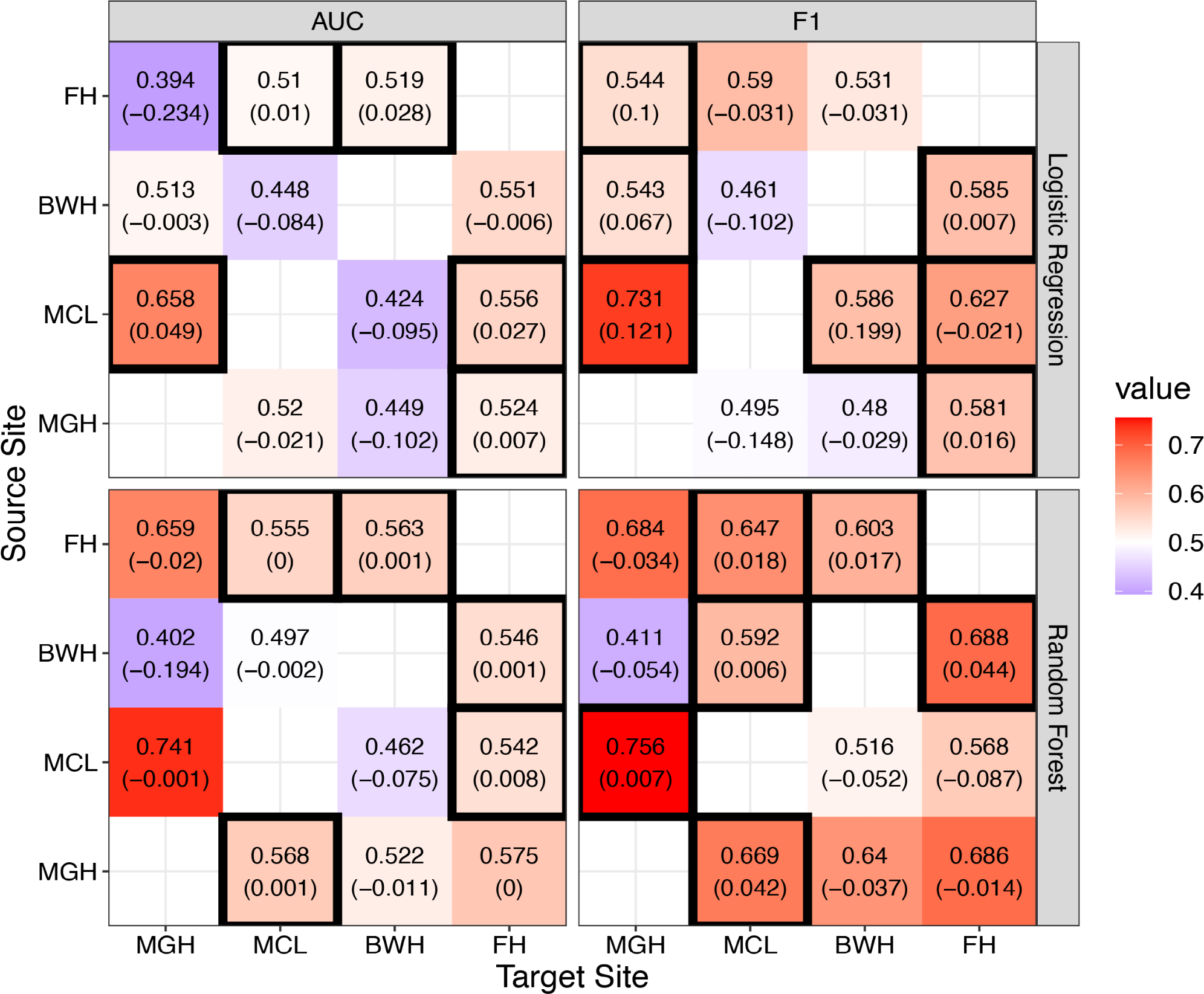
Cross-site prediction performance of the logistic regression models (top) and RF (bottom) after adjusting for covariate shift. The values outside the parentheses indicate the performance of the model with covariate shift adjustment while the values in the parentheses are the difference of the prediction performance of the adjusted model to the unadjusted model. Tiles with solid black borders are the cross-site predictions that benefit from covariate shift adjustment based on the change in the corresponding prediction performance metrics (AUC or F1).

## Discussion

This study focused on assessing characteristics of patient populations across four hospitals, performing cross-dataset validation, and comparing two prediction algorithms, logistic regression and RF, across independent hospital datasets. Our results showed that the distribution of the predictors differ across all sites and that prediction of readmission risk using structured EHR data is a very challenging task. Our results go further in establishing that there is a high amount of variability in results across sites, both in predictability and in the importance of different input variables, even when all four sites are part of the same health system, with shared databases, and in the same geographical area. Both models showed that LOS is an important feature of readmission risk. In the logistic regression model LOS is significantly associated with the outcome in three sites (Table 2) and in the RF model it has the highest importance among all predictors (Table 3). Longer LOS predicts greater readmission risk in BGW and Faulkner hospitals. The unexpected negative association in MGH might be explained by a proportion of patients with less severity and responding to hospital treatment in a short period, while the other patients with more severe symptoms needing longer hospitalized treatment, resulting in a “u shaped” association.

In addition, we showed that the most straightforward explanation for the lack of cross-site transportability, covariate shift, does not account for much of the performance difference. While there were significant differences in input variables distributions between sites, the negligible or even negative effect of our inverse probability weighting strategy suggests that more complex modes of heterogeneity across sites are present. For example, patients’ non-mental health related comorbidities may interact with mental health conditions in complex ways resulting in different conditional distributions in each site. As a result, modeling covariate shift using a simple reweighting strategy in this study is not able to capture a complex heterogeneity structure. The sites vary in multiple ways: The sample sizes McL and MGH are significantly bigger than BWH and Faulkers hospitals. Also, McL is a dedicated psychiatric facility while for the other three sites, psychiatry is just one of many specialties. It is currently unknown to what extent the diagnostic input codes from EHR relate to the readmission risk in a different way at different sites due to the heterogeneity of the patient profiles linked to different sites – for example, perhaps a site like McL is seeing patients with more complex medical and psychiatric comorbidities.

This study focused on the use of structured variables from EHRs. The unstructured data are richly detailed adding valuable contextual information to individual clinical records. It is likely that more fine-grained variables that can be extracted from unstructured text will yield higher predictive value. For example, for the hypothesis above concerning severity and comorbidities, an NLP-based approach mentioned may contribute – classifying psychiatric patients with or without non-mental health related comorbidities and modeling severity. Ongoing work is pursuing this direction.

Finally, we noted that the lack of transportability in predicting readmission seems to be less prominent between McLean and MGH—when applying the site-specific RF model of either hospital to the other, the resulting performance is comparable to those of the in-site predictions. One possible explanation for this anomalous result, as indicated by the feature importance in Table 2, is that the RF models for both hospitals share the same dominating predictor, LOS, which accounts for a non-ignorable proportion of the outcome variability. It is likely that there are unobserved yet highly predictive variables that are unique to each site and by including them in the models, the in-site predictions could be improved such that the anomaly we observed here for McLean and MGH is no longer present. This again highlights the necessity to move beyond the current set of structured variables for predictions of readmission.

## Data Availability

The data is not publicly available.

## Appendix A: Formal description of inverse probability weighting algorithm

We first introduce some notation for ease of exposition. Let (*Y, X, S*) be the triplet indicating the outcome, set of predictors and site index. Denote the loss function of a particular model by *l*(*Y*; *f*(*X*)), where *f* is the prediction function, and the distribution of (*Y, X*) in site *S* = *s* by *p*_*s*_. Under the assumption of covariate shift, we have *p*_*s*_(*y*|*x*) = *p*_*s*′_(*y*|*x*) but *p*_*s*_(*x*) ≠ *p*_*s*′_(*x*). The site-specific prediction model for site *s* is trained to optimize the expected loss 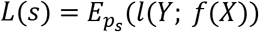. In practice, since *p*_*s*_ is unknown, the empirical loss 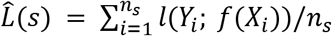 which is a consistent estimator of *L*(*s*), is used for training. Here *n*_*s*_ is the sample size of the training set for site *s*. If we want to build prediction models for site *s*′ based on data of site *s*, we can leverage the following result:

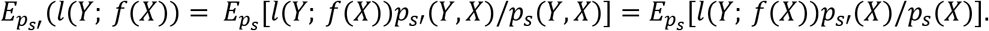

The last equality follows due to the covariate shift assumption. In other words, this means that we can use the following weighted empirical loss function, based on data in site *s*, to train a prediction function that asymptotically minimizes the expected loss for site *s*′:

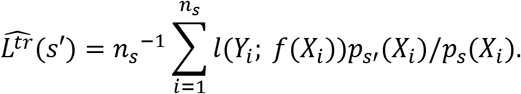

Note that the ratio *p*_*s*′_(*X*_*i*_)/*p*_*s*_(*X*) can be estimated using Bayes’ rule:

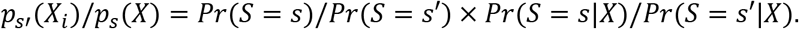

We estimate the first ratio to the right-hand side by *n*_*s*_/*n*_*s*′_ and the second ratio through logistic regression with the binary outcome *I*(*S* = *s*) and covariates *X* on the merged data of site *s* and *s*′.

